# Genome-wide meta-analysis of pilonidal sinus disease and its genetic overlap with hidradenitis suppurativa, hair patterning and adiposity

**DOI:** 10.64898/2026.10.31.26366973

**Authors:** João V. F. Cavalcante, Julia A. de Amorim, Rodrigo J. S. Dalmolin

**Author notes:** Corresponding author: João V. F. Cavalcante.

## Abstract

Pilonidal sinus disease (PSD) is a common, recurrent inflammatory disease of the natal cleft that clusters in families, but few genetic risk loci are known and its relationship with hidradenitis suppurativa (HS), acne and obesity is unclear. We meta-analysed public genome-wide summary statistics from the Million Veteran Program, UK Biobank, FinnGen and Genes & Health (7,868 cases and 1.6 million controls from four ancestry groups). Of 25 genome-wide significant loci outside the MHC, 19 were novel and 21 remained significant after correction for test-statistic inflation. Liability-scale SNP heritability was 13-15%. Fine-mapping resolved the *SOX2*, *PDE9A* and *KLF5* loci to single likely causal variants and identified a *WNT10A* missense variant, p.(Phe228Ile), associated with reduced risk. PSD was strongly genetically correlated with HS (rg = 0.76). In genomic structural equation models, the genetic correlation between PSD and body mass index (BMI, rg = 0.33) disappeared once HS was modelled (β = 0.01), whereas an independent component shared with male-pattern balding remained. Local genetic correlation and colocalisation located this overlap: six loci, including the *KLF5* and *SOX9* regions, shared a causal variant with HS and five with balding, whereas loci shared with BMI harboured distinct causal variants. Expression quantitative trait locus colocalisation implicated *SEMA4B* in skin and *WNT16* in fibroblasts. PSD is therefore a heritable disorder of the hair follicle that shares most of its common-variant liability with HS, with a separate hair-patterning component and a probable indirect association with obesity.

## Introduction

Pilonidal sinus disease (PSD) is a chronic inflammatory condition of the natal cleft in which hair-containing sinus tracts form and become repeatedly infected. ^1^ Its annual incidence has been estimated at 26 per 100,000 inhabitants. ^1^ It is more than twice as common in men, typically presents around the age of 20 years, and frequently recurs after surgery, particularly after midline closure. ^1,2^ PSD has traditionally been regarded as an acquired condition in which loose hairs are driven by friction into the skin of a deep, narrow natal cleft, particularly where the skin is softened or macerated. ^3^ However, 38% of patients report a family history of the disease, and familial cases present earlier and recur more often, ^1,4^ indicating that inherited factors also contribute.

Clinically, PSD is grouped with hidradenitis suppurativa (HS), acne conglobata and dissecting cellulitis of the scalp in the follicular occlusion tetrad, a set of conditions thought to share a pathological process initiated by follicular occlusion. ^5^ Whether this grouping reflects shared biology has not been established. The only dedicated genome-wide association study (GWAS) of PSD to date reported five risk loci and a genetic correlation with male-pattern baldness, ^6^ and biobank GWAS have released a few further associations, mostly with rare variants or below genome-wide significance (GWAS Catalog GCST90480439 and GCST90473908). ^7–9^ Larger GWAS of HS have identified loci near *SOX9* and *KLF5* that implicate the follicular epithelium, as well as variants that disrupt Notch and Wnt signalling. ^10–13^ Obesity is the most frequently cited risk factor for both diseases, but whether it influences PSD directly or through a liability shared with HS is unknown.

Several biobanks now release GWAS summary statistics for PSD. Each includes a few thousand cases at most, too few to detect most common risk variants individually, but together they provide substantially greater power, and analyses based on them can be reproduced entirely from public resources. Therefore, we combined five public datasets in the largest PSD GWAS meta-analysis to date in order to identify genetic risk loci for PSD, to determine how much of its genetic architecture is shared with HS, acne, hair patterning and adiposity and at which loci, and to prioritise candidate genes and tissues.

## Materials and methods

### Study design and data

This study used only publicly released GWAS summary statistics, and no individual-level data were accessed. The PSD cohorts were the Million Veteran Program (MVP, multi-ancestry and European-ancestry analyses), ^7^ UK Biobank whole-genome sequencing (European ancestry), ^8^ FinnGen release 13 (European ancestry) ^14^ and Genes & Health (British South Asian ancestry). ^15,16^ Summary statistics were obtained from the GWAS Catalog ^17^ and from the FinnGen public release. Case definitions were those of the contributing studies (Table S1). In UK Biobank and Genes & Health, cases had ICD-10 code L05 (pilonidal cyst or sinus) in linked health records. In FinnGen, cases had ICD-10 code L05 or ICD-9 code 685 in the hospital discharge or cause-of-death registries, and individuals with other skin infections were excluded from the controls, mapped to endpoint L12_PILONIDALCYST. In MVP, cases had at least two ICD-9 685 or ICD-10-CM L05 codes, mapped to phecode 686.3. Each cohort tested additive allelic effects using mixed or whole-genome regression models (SAIGE or REGENIE), which account for relatedness, with adjustment for age, sex and genetic principal components. Case definitions, genotyping or sequencing platforms, ancestry assignment and covariates for each cohort are listed in Table S1.

We performed two meta-analyses. META_ALL combined the MVP multi-ancestry analysis, UK Biobank, FinnGen and Genes & Health (7,868 cases and 1,598,492 controls), and META_EUR combined the MVP European-ancestry analysis, UK Biobank and FinnGen (6,771 cases and 1,374,212 controls). Because the MVP European-ancestry sample is contained within the MVP multi-ancestry analysis, the two were never combined. Analyses that depend on a European LD reference (LDSC, Genomic SEM, LAVA and SuSiE) used META_EUR, because 86% of cases were of European ancestry and a European LD reference does not represent a mixed-ancestry sample. Heritability, genetic correlations and colocalisation were also estimated in META_ALL as a sensitivity analysis. Comparator traits were HS, from a meta-analysis of MVP, UK Biobank and FinnGen (3,941 cases, 1,077 of them of African ancestry, and 1,435,603 controls, GWAS Catalog GCST90726609), ^13^ and HS in FinnGen alone (ICD-10 L73.2, 1,453 cases). We also used acne, in FinnGen (ICD-10 L70, 4,812 cases) and in a meta-analysis of three Europeanancestry cohorts (34,422 cases), ^18^ alopecia areata in FinnGen (ICD-10 L63, 994 cases), hair pattern 1 (“no balding”, the first of four patterns reported by UK Biobank men, 66,172 of 207,036 men), ^19^ and body mass index (BMI) and waist-to-hip ratio adjusted for BMI (WHRadjBMI) from GIANT and UK Biobank. ^20^ The FinnGen comparator endpoints shared 474,896 controls, from which individuals with disorders of the skin appendages were excluded.

### Harmonisation and meta-analysis

All PSD summary statistics were released on GRCh38 and were combined by chromosome, position and alleles. We retained autosomal variants with valid, distinct alleles and non-missing effect sizes, harmonised allele order and effect direction across cohorts. Imputation quality filters were those applied before release, imputation quality > 0.3 in MVP ^7^ and ≥ 0.6 in FinnGen, ^14^ and the UK Biobank and Genes & Health data were from genome and exome sequencing. Within each cohort we retained variants with a minor allele frequency of at least 1%, removed exact duplicates (same position and alleles) and excluded multi-allelic sites. Cohorts were combined by fixed-effect inverse-variance-weighted meta-analysis using METAL tool (standarderror scheme), ^21^ and between-cohort heterogeneity was quantified with Cochran’s Q and the I² statistic using METAL’s ANALYZE HETEROGENEITY option. Variants were considered for discovery if they were present in at least two cohorts and reached *p* < 5×10^−8^. Lead variants were selected by distance (±500 kb), and loci from the two meta-analyses were merged into a single list. Signals in the MHC (chr6:25-34 Mb) were reported separately and excluded from downstream locus-level analyses because of the region’s extended linkage disequilibrium (LD). Loci within 500 kb of a previously published PSD association ^6–9,22^ were classified as known and all others as novel.

We assessed robustness in three ways. We rescaled test statistics by the LD score regression (LDSC) intercept of the corresponding meta-analysis (χ²/intercept), re-estimated lead-variant effects without FinnGen, the largest cohort, and fitted DerSimonian-Laird random-effects models with the metafor R package ^23,24^ at each lead variant.

### Fine-mapping and variant annotation

For each lead variant, we computed approximate Bayes factors and 95% credible sets within ±500 kb in both meta-analyses with the finemap.abf function of the coloc R package, which assumes a single causal variant per locus. It uses a standard deviation of 0.2, which corresponds to 95% prior belief that the relative risk is below 1.5. ^25,26^ For META_EUR, we additionally ran SuSiE-RSS ^27^ (up to 10 signals, 95% coverage, minimum purity 0.5) using UK Biobank European LD matrices, ^28^ with LD-mismatch diagnostics (the estimated LD-consistency parameter and outliers in the conditional-distribution diagnostic implemented in susieR). Credible-set variants were annotated with the Ensembl Variant Effect Predictor. ^29^

### Heritability and genetic correlation

SNP heritability, the LDSC intercept and genetic correlations (rg) were estimated with LDSC using HapMap3 variants and European LD scores. ^30,31^ Observed-scale heritability was converted to the liability scale ^32^ assuming population prevalences of 0.5% and 1%, which span the proportion of cases in the contributing cohorts (0.3-1.0%, Table S1). Using Genomic SEM ^33^ with diagonally weighted least squares estimation, we fitted a common-factor model of PSD, HS and FinnGen acne, and multivariable regressions of PSD on HS and BMI and on HS, acne, BMI, WHRadjBMI and balding jointly. Sample overlap between traits was accounted for through the LDSC cross-trait intercepts.

### Local genetic correlation and colocalisation

Local genetic correlations were estimated with LAVA ^34^ across 2,495 LD-independent blocks, using the UK Biobank European reference panel and correcting for sample overlap with the LDSC cross-trait intercepts. Bivariate tests were performed in non-MHC blocks in which both traits had univariate local heritability at *p* < 10^−4^, and multiple testing was controlled with the Benjamini-Hochberg false discovery rate. To test for shared causal variants, we applied coloc.abf ^35,36^ in ±500 kb windows around each PSD locus and each genome-wide significant HS locus, with priors *p*_1_ = *p*_2_ = 10^−4^ and *p*_12_ = 5×10^−6^. Variants were matched by rsID, and at least 50 shared variants were required. A shared causal variant was inferred when the posterior probability PP.H4 exceeded 0.8.

### Candidate effector genes

Each non-MHC locus was colocalised, with the same priors, with GTEx v8 cis-eQTLs ^37^ from the eQTL Catalogue ^38^ in sun-exposed and non-sun-exposed skin, cultured fibroblasts, and subcutaneous and visceral adipose tissue. Genes with an eQTL at *p* < 10^−4^ in the window were tested. Gene-level association was computed with MAGMA. ^39^ For gene-set analysis, genes in the MHC were removed from the gene universe and MAGMA competitive tests were run on 6,717 Gene Ontology biological process and Reactome gene sets of 10-500 genes from MSigDB 2025.1, with Bonferroni correction across sets. For significant sets, we repeated the test after removing genes within 1 Mb of the most associated member gene and after removing genome-wide significant genes.

Genes near each locus were then scored on five lines of evidence: proximity (nearest gene), the top MAGMA gene within 250 kb (*p* < 10^−3^), a coding variant in the credible set (posterior inclusion probability [PIP] ≥ 0.01, with stronger weight for PIP ≥ 0.1), eQTL colocalisation (PP.H4 0.5-0.8, with stronger weight for PP.H4 > 0.8) and the top gene from the Open Targets locus-to-gene model. ^40^ A protein-coding gene with functional evidence (a coding variant with PIP ≥ 0.1 or eQTL colocalisation with PP.H4 > 0.8) was prioritised over proximity.

### Use of a large language model

A large language model (Claude, Anthropic) assisted with writing analysis code. The authors reviewed all code, checked all reported results against the analysis outputs, and take full responsibility for the content.

## Results

### Twenty-five risk loci for PSD, 19 of them novel

Both meta-analyses showed moderate genomic inflation (λ_GC_ 1.13 for META_EUR and 1.11 for META_ALL, Fig. S1). The LDSC intercept was 1.05 and the attenuation ratio 0.25, indicating that approximately three quarters of the inflation reflects polygenic signal rather than confounding. We identified 25 genome-wide significant loci outside the MHC, together with an MHC signal at *HLA-DQA1* (rs1281932) that was not analysed further (Fig. 1, Table 1). Six loci had been reported previously (Table S3). Four of them (*PRKAR1B*, *TMED10/NEK9*, *TBX15* and *HDAC9/TWIST1*) overlapped loci reported by Roberson et al., ^6^ and two (*DKK1/MBL2* and *S100A11/TCHH*) were genome-wide significant in FinnGen release 12. ^22^ Of the remaining 19, none reached genome-wide significance in any single cohort.

**Figure 1.**
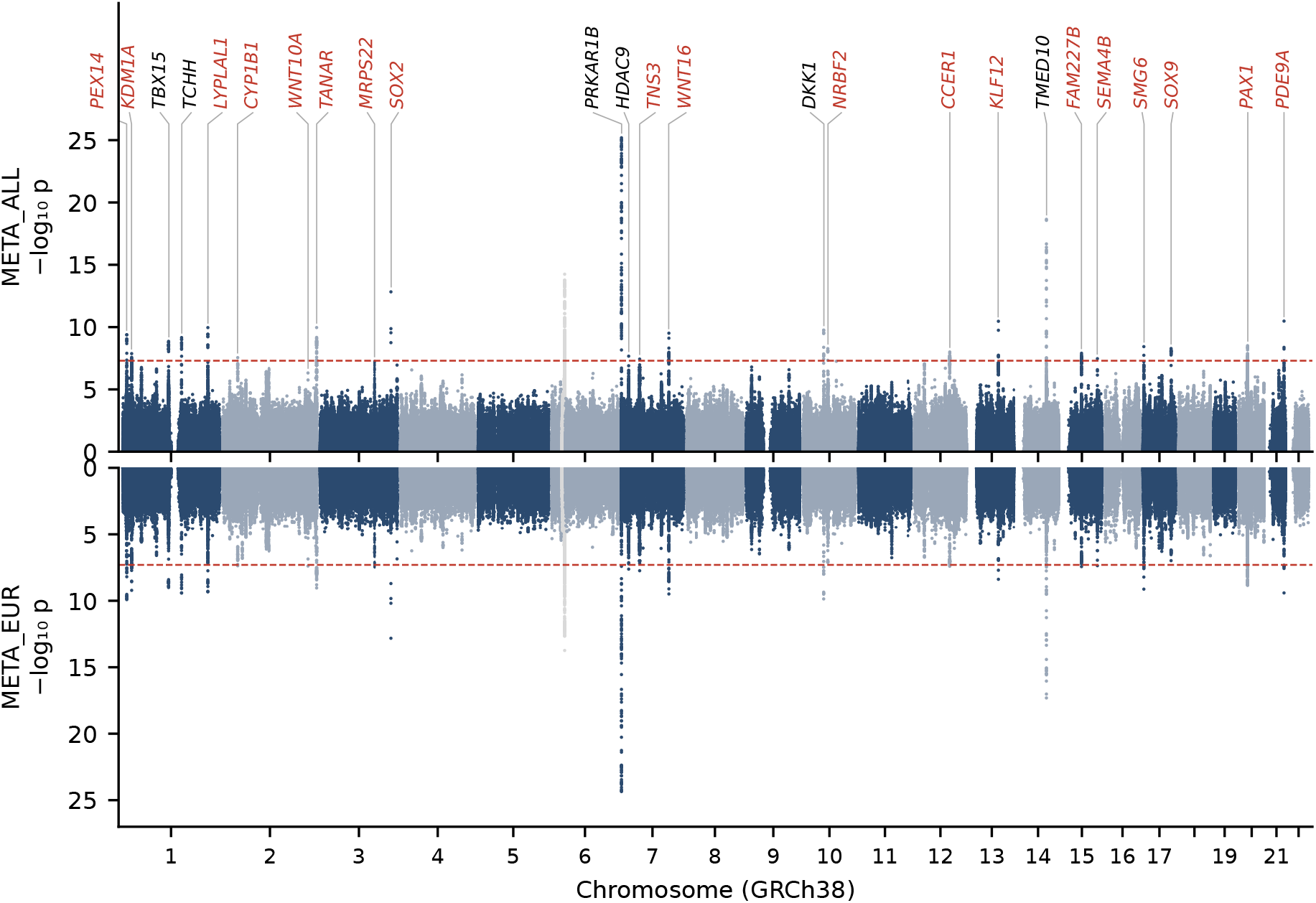
Genome-wide meta-analysis of pilonidal sinus disease. Miami plot of the multiancestry (META_ALL, top) and European (META_EUR, bottom) meta-analyses. Variants present in at least two cohorts are shown (GRCh38), and the dashed line marks *p* = 5×10^−8^. The 25 non-MHC loci are labelled with their prioritised or nearest gene (novel loci in red and previously reported loci in black). The MHC is shown in grey.

**Table 1:** Twenty-five genome-wide significant pilonidal sinus disease loci outside the MHC.

| Locus (GRCh38) | Lead variant | EA/OA | EAF | OR (95% CI), EUR | *p*, EUR | OR (95% CI), ALL | *p*, ALL | Status | Nearest gene | Prioritised gene |
|---|---|---|---|---|---|---|---|---|---|---|
| 1:10,598,164 | rs284265 | C/T | 0.15 | 1.163 (1.111-1.218) | 1.2×10<sup>−10</sup> | 1.148 (1.099-1.198) | 4.5×10<sup>−10</sup> | novel | *PEX14* | *PEX14* (lncRNA) |
| 1:23,080,604 | rs579098 | A/G | 0.12 | 1.176 (1.117-1.238) | 5.9×10<sup>−10</sup> | 1.150 (1.096-1.207) | 1.4×10<sup>−8</sup> | novel | *KDM1A* | *KDM1A* |
| 1:118,947,259 | rs10923703 | C/T | 0.56 | 1.115 (1.077-1.155) | 9.6×10<sup>−10</sup> | 1.111 (1.074-1.149) | 1.5×10<sup>−9</sup> | known | *TBX15* | *TBX15* |
| 1:152,032,328 | rs12123975 | A/G | 0.74 | 0.885 (0.851-0.919) | 3.7×10<sup>−10</sup> | 0.894 (0.861-0.927) | 2.1×10<sup>−9</sup> | known | *S100A11* | *TCHH* |
| 1:219,024,094 | rs141106808 | G/GA | 0.86 | 0.861 (0.820-0.903) | 1.1×10<sup>−9</sup> | 0.858 (0.819-0.899) | 1.2×10<sup>−10</sup> | novel | *LYPLAL1* | *LYPLAL1* |
| 2:38,089,429 | rs151096<sup>†</sup> | A/C | 0.54 | 1.101 (1.064-1.140) | 4.1×10<sup>−8</sup> | 1.100 (1.064-1.138) | 3.0×10<sup>−8</sup> | novel | *CYP1B1* | *CYP1B1* |
| 2:218,890,289 | rs121908120<sup>†‡</sup> | A/T | 0.02 | 0.540 (0.434-0.673) | 3.9×10<sup>−8</sup> | 0.580 (0.470-0.718) | 5.0×10<sup>−7</sup> | novel | *WNT10A* | *WNT10A* |
| 2:240,936,015 | rs56155367<sup>‡</sup> | G/GA | 0.33 | 1.135 (1.090-1.182) | 8.9×10<sup>−10</sup> | 1.137 (1.093-1.182) | 1.2×10<sup>−10</sup> | novel | *CROCC2* | *TANAR* (lncRNA) |
| 3:139,120,151 | rs940190<sup>†</sup> | A/C | 0.22 | 1.121 (1.077-1.168) | 3.3×10<sup>−8</sup> | 1.110 (1.068-1.154) | 1.1×10<sup>−7</sup> | novel | *MRPS22* | *MRPS22*/*PRR23B* |
| 3:181,794,163 | rs1345417 | C/G | 0.37 | 0.870 (0.839-0.903) | 1.4×10<sup>−13</sup> | 0.875 (0.844-0.906) | 1.5×10<sup>−13</sup> | novel | *SOX2* | *SOX2* |
| 7:543,758 | rs35369699 | C/T | 0.86 | 0.780 (0.744-0.818) | 4.2×10<sup>−25</sup> | 0.781 (0.746-0.818) | 6.8×10<sup>−26</sup> | known | *PRKAR1B* | *PRKAR1B* |
| 7:18,874,234 | rs35536284<sup>‡</sup> | G/GT | 0.64 | 0.893 (0.859-0.929) | 2.3×10<sup>−8</sup> | 0.893 (0.859-0.929) | 2.3×10<sup>−8</sup> | known | *HDAC9* | *HDAC9* |
| 7:46,848,844 | rs10951876 | A/C | 0.49 | 1.103 (1.066-1.141) | 1.7×10<sup>−8</sup> | 1.096 (1.061-1.133) | 3.9×10<sup>−8</sup> | novel | *TNS3* | *TNS3* |
| 7:121,300,382 | rs2707521 | C/T | 0.66 | 0.893 (0.861-0.925) | 3.1×10<sup>−10</sup> | 0.895 (0.865-0.927) | 3.3×10<sup>−10</sup> | novel | *CPED1* | *WNT16* |
| 10:52,574,893 | rs7905367 | C/G | 0.74 | 1.148 (1.101-1.198) | 1.3×10<sup>−10</sup> | 1.139 (1.094-1.186) | 3.6×10<sup>−10</sup> | known | *MBL2* | *DKK1*/*MBL2* |
| 10:63,085,128 | rs72824926 | A/G | 0.76 | 0.897 (0.863-0.933) | 7.2×10<sup>−8</sup> | 0.891 (0.856-0.926) | 5.7×10<sup>−9</sup> | novel | *NRBF2* | *NRBF2* |
| 12:90,786,242 | rs2089111 | C/G | 0.75 | 0.896 (0.862-0.932) | 4.4×10<sup>−8</sup> | 0.894 (0.860-0.929) | 1.1×10<sup>−8</sup> | novel | *CCER1* | *CCER1* |
| 13:73,432,270 | rs17090189 | A/G | 0.86 | 1.163 (1.106-1.223) | 3.9×10<sup>−9</sup> | 1.175 (1.120-1.233) | 3.8×10<sup>−11</sup> | novel | *KLF12* | *KLF12* |
| 14:75,226,898 | rs73313647 | A/T | 0.77 | 0.840 (0.807-0.874) | 4.7×10<sup>−18</sup> | 0.840 (0.808-0.872) | 2.5×10<sup>−19</sup> | known | *TMED10* | *TMED10* |
| 15:49,715,064 | rs17483319 | G/T | 0.34 | 1.107 (1.068-1.148) | 3.5×10<sup>−8</sup> | 1.108 (1.070-1.148) | 1.3×10<sup>−8</sup> | novel | *DTWD1* | *FAM227B* |
| 15:90,210,523 | rs1399238<sup>†</sup> | A/G | 0.79 | 1.130 (1.081-1.180) | 4.1×10<sup>−8</sup> | 1.126 (1.079-1.174) | 3.6×10<sup>−8</sup> | novel | *SEMA4B* | *SEMA4B* |
| 17:2,152,586 | rs11650850 | A/C | 0.38 | 0.895 (0.863-0.927) | 7.3×10<sup>−10</sup> | 0.902 (0.872-0.934) | 3.9×10<sup>−9</sup> | novel | *SMG6* | *SMG6* (lncRNA) |
| 17:71,519,473 | rs17226067 | A/G | 0.91 | 0.861 (0.813-0.912) | 2.8×10<sup>−7</sup> | 0.856 (0.812-0.902) | 5.6×10<sup>−9</sup> | novel | *SOX9* | *SOX9* |
| 20:22,109,943 | rs201150 | A/T | 0.63 | 1.117 (1.078-1.158) | 1.4×10<sup>−9</sup> | 1.106 (1.068-1.145) | 1.4×10<sup>−8</sup> | novel | *PAX1* | *PAX1* |
| 21:42,644,024 | rs111363985 | G/T | 0.94 | 0.811 (0.759-0.866) | 3.8×10<sup>−10</sup> | 0.804 (0.753-0.858) | 3.6×10<sup>−11</sup> | novel | *PDE9A* | *PDE9A* |

EA/OA, effect/other allele. EAF, effect-allele frequency in META\_EUR. OR, odds ratio per effect allele from fixed-effect inverse-variance meta-analysis (METAL). EUR, European meta-analysis (MVP European, UK Biobank, FinnGen). ALL, multi-ancestry meta-analysis (MVP multi-ancestry, UK Biobank, FinnGen, Genes & Health). Prioritised gene: locus-to-gene scoring (Materials and Methods). <sup>†</sup> $5 \times 10^{-8} < p < 10^{-7}$ after LDSC-intercept correction. <sup>‡</sup> Lead variant measured in two of the three European cohorts.

Several analyses supported the robustness of these associations. All 19 novel loci showed a consistent direction of effect, with *p* < 0.05, in the cohorts other than FinnGen (Table S4). After rescaling by the LDSC intercept, 21 loci, 15 of them novel, remained genome-wide significant, whereas the *CYP1B1*, *SEMA4B*, *MRPS22* and *WNT10A* loci fell just short of the threshold (*p* = 5-8×10^−8^, Table S4). Twenty-one loci showed no significant between-cohort heterogeneity. The exceptions were *DKK1/MBL2*, *S100A11/TCHH*, *KDM1A* and *HDAC9* (I² 67-81%, Table S2, Fig. S2). At these loci, the MVP estimate differed in magnitude from those of the other cohorts or, at *HDAC9*, the lead indel variant was measured in only two cohorts.

In META_EUR, liability-scale SNP heritability was 13.1% (SE 1.2%) assuming a prevalence of 0.5% and 15.3% (SE 1.4%) assuming 1% (Table S7). The genetic correlation between PSD in MVP and in FinnGen was 0.83 (SE 0.18) (Table S8), consistent with a largely shared genetic basis across biobanks despite differences in case ascertainment.

### Fine-mapping resolves several loci to single variants

At the *SOX2* locus, a single variant, rs1345417, accounted for almost all of the posterior probability (PIP 0.99-1.00, Table S5) and formed a one-variant credible set. Fine-mapping also resolved the *PDE9A* locus (rs111363985, PIP 0.90-0.98) and the *KLF5* region on 13q22 (rs17090189, PIP 0.65-0.81). This variant is also the lead variant reported for HS at this locus by Sun et al. ^10^ At *WNT10A*, the credible set contained a low-frequency missense variant, rs121908120 (NM_025216.3:c.682T>A, p.(Phe228Ile), effect allele frequency 2.4%, approximate Bayes factor PIP 0.45-0.57, SuSiE PIP 0.73, Table S6), whose minor allele was associated with reduced risk (odds ratio 0.54, 95% CI 0.43-0.67). The same allele is a recognised cause of *WNT10A*-related ectodermal dysplasia (ClinVar variation 4462) and has been associated with acne. ^41^

### PSD shares most of its common-variant liability with HS

PSD was genetically correlated with HS (rg = 0.76, SE 0.07, *p* = 7×10^−25^), with acne (rg = 0.43, SE 0.08, *p* = 6×10^−8^ with FinnGen acne and rg = 0.15, SE 0.05, *p* = 0.003 with the Teder-Laving et al. GWAS) and with BMI (rg = 0.33, SE 0.03, *p* = 2×10^−25^) (Fig. 2a, Table S8). HS was itself genetically correlated with BMI (rg = 0.42). Estimates from META_ALL were nearly identical (rg = 0.76 with HS and 0.33 with BMI). The correlation with HS persisted in non-overlapping samples (MVP PSD and FinnGen HS, rg = 0.54, SE 0.20). PSD was weakly correlated with balding (rg = −0.13 with the “no balding” pattern, *p* = 0.025) and was not correlated with WHRadjBMI or alopecia areata.

**Figure 2.**
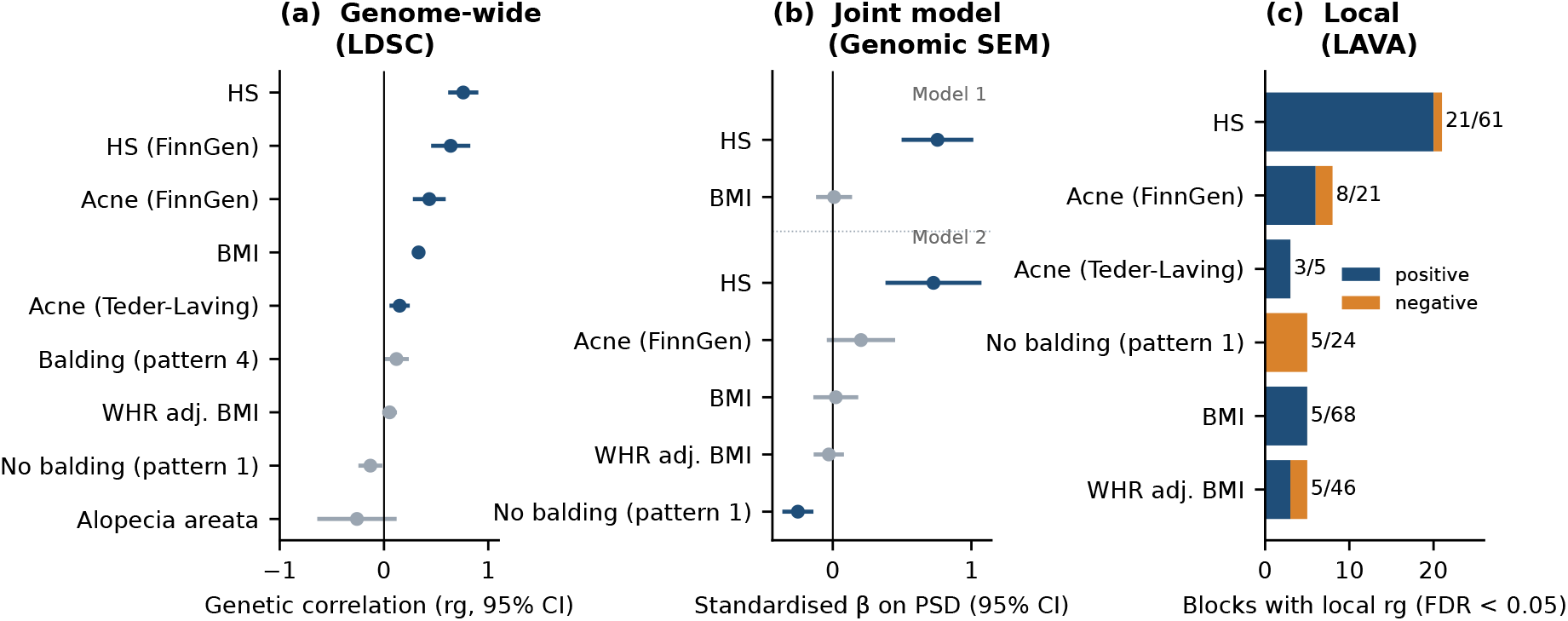
Genetic overlap of pilonidal sinus disease (PSD) with follicular occlusion, hairpatterning and adiposity traits. (a) Genome-wide genetic correlations from LD score regression (META_EUR) with 95% confidence intervals. Bonferroni-significant correlations are shown in blue. (b) Genomic structural equation modelling: standardised effects (95% confidence intervals) on PSD in a model with hidradenitis suppurativa (HS) and body mass index (BMI) (model 1) and with HS, acne, BMI, waist-to-hip ratio adjusted for BMI (WHR adj. BMI) and balding (model 2). (c) Number of LD blocks with significant local genetic correlation (LAVA, false discovery rate < 0.05) among blocks tested for each trait. “No balding” (pattern 1) is the reference pattern, so negative correlations indicate that PSD risk tracks balding.

Genomic SEM separated these overlapping correlations (Fig. 2b, Table S9). In a model of PSD on HS and BMI, HS was associated with PSD (standardised β = 0.75, SE 0.13, *p* = 1×10^−8^), whereas BMI was not (β = 0.01, *p* = 0.89). When acne, WHRadjBMI and balding were added, HS remained the main predictor (β = 0.73, *p* = 4×10^−5^), and an independent hair-patterning component emerged: the “no balding” pattern was negatively associated with PSD (β = −0.25, SE 0.06, *p* = 1×10^−5^). In a common-factor model of PSD, HS and FinnGen acne, PSD loaded at 0.94 (HS 0.81, acne 0.47), leaving little PSD-specific residual variance (0.12, SE 0.26).

### The overlap with HS and balding maps to specific loci and variants

Local genetic correlation analysis located the shared liability within the genome (Fig. 2c, Table S11). Of the 101 non-MHC blocks with local PSD heritability, 61 could be tested with HS, and 21 of these showed significant local genetic correlation (false discovery rate < 0.05), 20 of them positive. All five significant blocks with the “no balding” pattern (of 24 tested) showed negative correlations, indicating that PSD risk also tracked balding at the local level. In contrast, local genetic correlation with BMI was close to zero in 15 of the 16 testable blocks with nominal (*p* < 0.05) local correlation with HS.

Colocalisation resolved these overlaps to individual variants (Fig. 3, Table S10). Six PSD loci shared a causal variant with HS (PP.H4 > 0.8): *PRKAR1B*, *LYPLAL1*, *CYP1B1*, *WNT10A*, and the *KLF5* and *SOX9* regions. Three of these also colocalised with the FinnGen-only HS GWAS. For acne, *SEMA4B* colocalised with both acne GWAS and *LYPLAL1* with FinnGen acne. Five loci shared a causal variant with balding: *HDAC9/TWIST1*, *DKK1/MBL2*, *S100A11/TCHH*, *SOX2*, and the *PAX1* region on 20p11. At three of these (*HDAC9/TWIST1*, *S100A11/TCHH* and 20p11) the PSD risk allele was associated with increased balding, and at the other two (*DKK1/MBL2* and *SOX2*) with decreased balding. No PSD locus shared a causal variant with BMI. At nine loci with association signals for both traits, the posterior probabilities favoured distinct causal variants, and only *WNT10A* colocalised with WHRadjBMI. With META_ALL, the same six loci colocalised with HS, and four of the five balding loci colocalised with balding (PP.H4 = 0.61 at 20p11).

**Figure 3.**
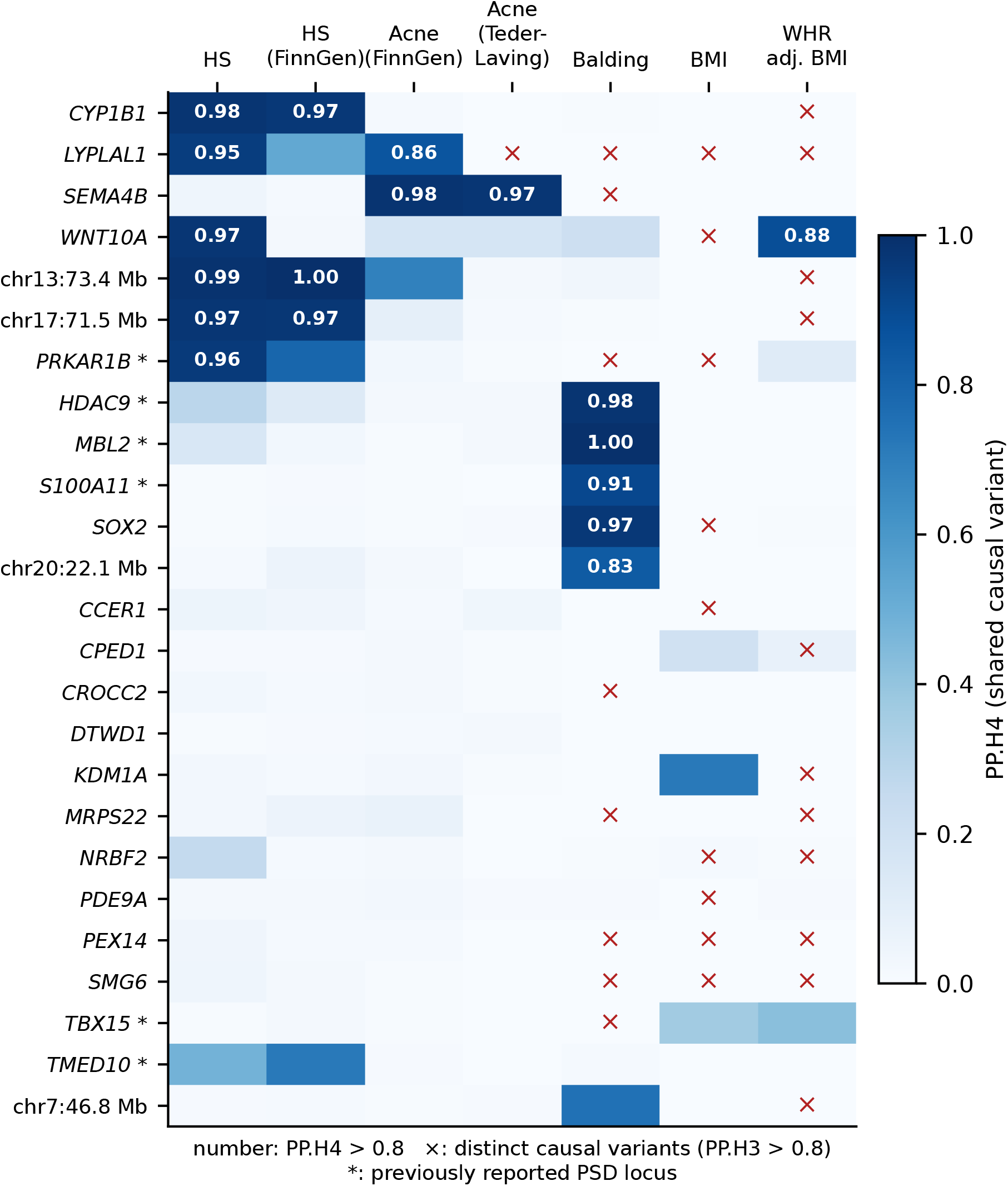
Shared causal variants between pilonidal sinus disease (PSD) loci and related traits. Posterior probability of a shared causal variant (coloc PP.H4) at each non-MHC PSD locus (META_EUR, ±500 kb). Numbers are shown for PP.H4 > 0.8, and × marks distinct causal variants (PP.H3 > 0.8). Asterisks mark previously reported PSD loci.

### Candidate effector genes in skin

Six loci colocalised with an eQTL (PP.H4 > 0.8, Fig. 4a, Table S12). At *SEMA4B*, the PSD risk allele was associated with increased *SEMA4B* expression in both skin tissues (PP.H4 0.97). The shared signal resolved to a single variant, rs34560261 (variant-level posterior probability 0.96-1.00), which lies in a p63-bound intronic element and is the likely causal variant for acne at this locus. ^41^ At the *CPED1* locus, the risk allele was associated with increased *WNT16* expression in cultured fibroblasts (PP.H4 0.94) and subcutaneous adipose tissue (PP.H4 0.90), whereas *WNT16* expression in skin was driven by a different variant. At the four remaining loci, the colocalising gene was a long non-coding RNA (lncRNA): an antisense transcript of *PEX14* in fibroblasts and skin (PP.H4 0.95 and 0.84), *TANAR* at the *CROCC2* locus and *RMDN2-AS1* at the *CYP1B1* locus, both in visceral adipose tissue, and an antisense transcript of *SMG6* in subcutaneous adipose tissue.

**Figure 4.**
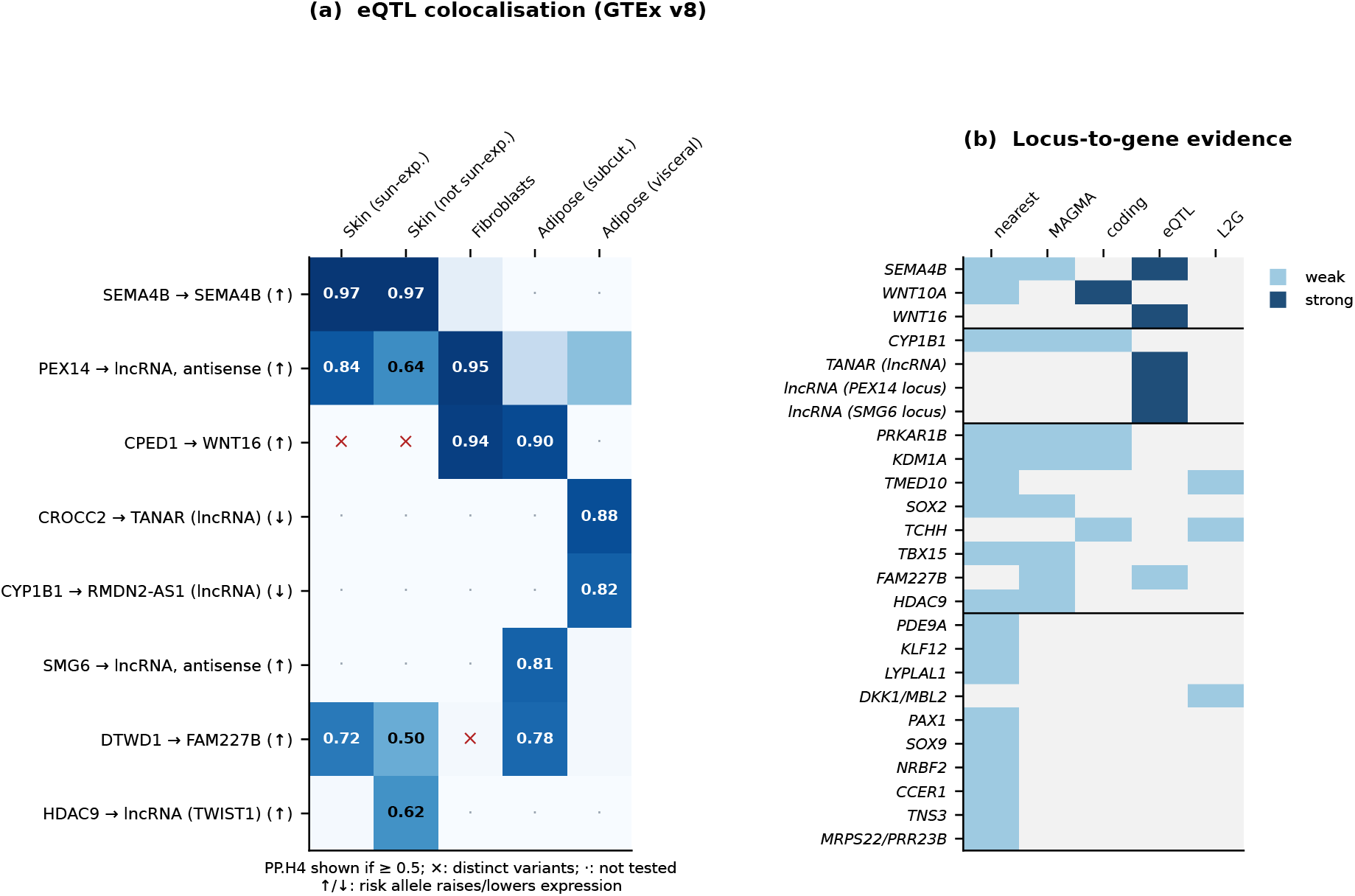
Candidate effector genes. (a) Colocalisation of pilonidal sinus disease (PSD) loci with GTEx v8 eQTLs in skin, fibroblasts and adipose tissue (PP.H4 shown if ≥ 0.5). Arrows indicate whether the PSD risk allele raises or lowers expression. (b) Locus-to-gene evidence for the prioritised gene at each locus, grouped by evidence tier: functional support for a protein-coding gene, functional support for a lncRNA only, two or more weaker lines, and one line.

Integrating all lines of evidence, three loci had a protein-coding candidate gene with functional support (*SEMA4B*, *WNT16* and *WNT10A*), four had functional support only for a lncRNA, eight had two or more weaker lines of evidence, and ten were assigned on proximity alone (Fig. 4b, Table S13). Several of the genes assigned on proximity regulate hair-follicle development: *SOX2* marks a dermal papilla subpopulation that specifies hair-follicle type ^42^ and *SOX9* the outer root sheath and stem-cell niche, ^43^ *DKK1* is induced by androgens in dermal papilla cells from balding scalp, ^44^ and *TCHH* encodes trichohyalin, a structural protein of the inner root sheath and of the hair-shaft medulla. ^45^ In a genome-wide gene-set analysis, one set passed correction for multiple testing, intermediate filament organisation (74 genes, β = 0.57, SE 0.13, *p* = 5×10^−6^, Bonferroni-corrected *p* = 0.03, META_ALL, Table S14). The association was driven by *TCHH* and by hair and inner root sheath keratins (*KRT31*, *KRT35*, *KRT36*, *KRT40* and *KRT71*), none of which reached genome-wide significance, and it persisted after removal of the *TCHH* region (*p* = 3×10^−5^). The set showed the same direction in META_EUR (*p* = 7×10^−5^), and it was not associated with HS, acne or BMI (*p* > 0.6).

Four loci, including the *SOX9* and *KLF5* regions, lie in gene deserts 250-600 kb from the nearest gene.

## Discussion

In the largest GWAS meta-analysis of PSD to date, we identified 25 risk loci, 19 of them novel, and found that the genetic architecture of PSD comprises two components. The larger is shared with HS, genome-wide, within specific genomic regions and at individual causal variants. The second is a hair-patterning component shared with male-pattern balding. Obesity, the most frequently cited risk factor, contributed no independent genetic signal.

The overlap with HS provides genetic support for the follicular occlusion concept. ^5^ The two diseases share most of their common-variant liability, with a genetic correlation of 0.76, and the shared signal is concentrated at loci implicating the follicular epithelium. The *SOX9* and *KLF5* regions are the strongest HS loci, ^10,12^ and HS risk alleles at both alter transcriptional activity in reporter assays. ^12^ At the *KLF5* region, the PSD lead variant is the HS lead variant reported by Sun et al., ^10^ although the primary signal in a larger HS GWAS was a different variant in low LD with it (rs981625, r^2^ = 0.24). ^12^ *SEMA4B* and *WNT10A* are acne loci whose causal variants act in keratinocyte regulatory elements or in the WNT pathway, ^41^ which is also disrupted by HS risk variants, ^11^ and the WNT16B isoform promotes keratinocyte proliferation. ^46^ Taken together, these findings suggest that inherited properties of the follicular epithelium, rather than immune dysregulation, initiate PSD. Outside the MHC, no immune pathway was enriched in gene-set analysis, which is consistent with the inflammation of PSD arising secondarily to follicle rupture and hair insertion, although genetic data alone cannot establish the order of these events.

The hair-patterning component distinguishes PSD from HS. Roberson et al. first reported a genetic correlation between PSD and balding. ^6^ We show that this correlation persists after accounting for HS and that it maps to five shared causal variants near regulators of hair-follicle development and of the hair shaft. Beyond the genome-wide significant loci, PSD risk was enriched in genes encoding structural proteins of the hair shaft and inner root sheath.

Genome-wide, and in every significant local block, PSD risk tracked balding. At the shared variants, however, effects were concordant at three loci and opposite at two (*DKK1/MBL2* and *SOX2*), suggesting that these genes influence the two traits through different mechanisms. These results are consistent with the long-standing view that hair is central to PSD pathogenesis. ^3^ Inherited, partly androgen-dependent properties of hair and hair follicles could make hairs more likely to penetrate the skin of the natal cleft and might contribute to the male predominance and post-pubertal onset of the disease.

Obesity is associated with PSD in clinical series, ^1^ and we observed a positive genetic correlation with BMI, as reported previously. ^6^ However, this correlation disappeared once HS was modelled, BMI showed no local genetic correlation in the regions shared with HS, and loci associated with both traits harboured distinct causal variants. A possible explanation is that the PSD-BMI correlation reflects pleiotropy shared with HS, which is itself genetically correlated with BMI, rather than a causal effect of adiposity on PSD. The clinical association between PSD and high BMI may nonetheless reflect local mechanical factors, such as a deeper natal cleft, friction and sweating, that are not captured by genetic liability to BMI.

Our results do not bear directly on treatment, but they agree with clinical observations. The surgical procedures with the lowest recurrence rates flatten the natal cleft and avoid a midline wound, ^2^ which fits a disease driven by hair penetrating the skin of the cleft. The shared genetic basis with HS is consistent with the reported co-occurrence of the two diseases. ^5^

However, there are a few limitations to our approach. Approximately 86% of PSD cases were of European ancestry. The HS comparator GWAS includes African-ancestry participants (27% of its cases), whom European LD scores do not represent exactly. Although the loci showed consistent directions of effect outside FinnGen, replication in an independent cohort is still required. Additionally, four novel loci fell just short of genome-wide significance after intercept correction. Finally, eQTL data from adult bulk tissues do not capture developmental gene regulation, which probably explains why the developmental genes at several loci lacked eQTL support.

In conclusion, PSD is a heritable disorder of the hair follicle that shares most of its commonvariant liability with HS and has a distinct hair-patterning component, whereas its association with obesity appears to be indirect. Future studies should test whether genetic effects differ by sex and age at onset, and characterise the candidate genes, particularly *SEMA4B*, *WNT16* and *WNT10A*, in the follicular epithelium.

## Data Availability

All input summary statistics are publicly available from the sources listed in Table S1. The meta-analysis summary statistics are available from Zenodo (https://doi.org/10.5281/zenodo.23835851).

## Code availability

All analysis code is available at https://github.com/dalmolingroup/pilonidal_genetics.

## Acknowledgements

We thank the participants and investigators of the Million Veteran Program, UK Biobank, FinnGen and Genes & Health, and the GWAS Catalog, GTEx and the eQTL Catalogue for making their data publicly available. We thank NPAD / UFRN for computational resources.

## Author contributions

JVFC: Conceptualization, Formal analysis, Investigation, Methodology, Visualization, Writing - original draft, Writing - review and editing. JAA: Methodology, Writing - review and editing. RJSD: Supervision, Writing - review and editing.

## Ethical approval

Ethical approval was not required. This study used only publicly released, de-identified genome-wide association summary statistics, and each contributing study obtained approval from its own ethics committee and informed consent from its participants.

