## Supplementary Figures S1 and S2 for "Genome-wide meta-analysis of pilonidal sinus disease and its genetic overlap with hidradenitis suppurativa, hair patterning and adiposity"

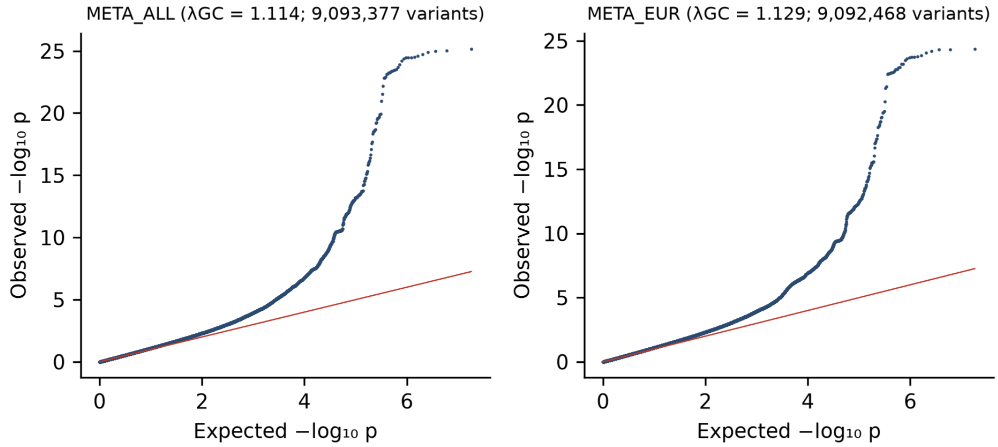


**Figure S1. Quantile-quantile plots** of the multi-ancestry (META_ALL) and European (META_EUR) meta-analyses, for variants present in at least two cohorts. $\lambda_{GC}$ is the genomic inflation factor. The LDSC intercept (1.05) shows that most of the inflation reflects polygenicity.

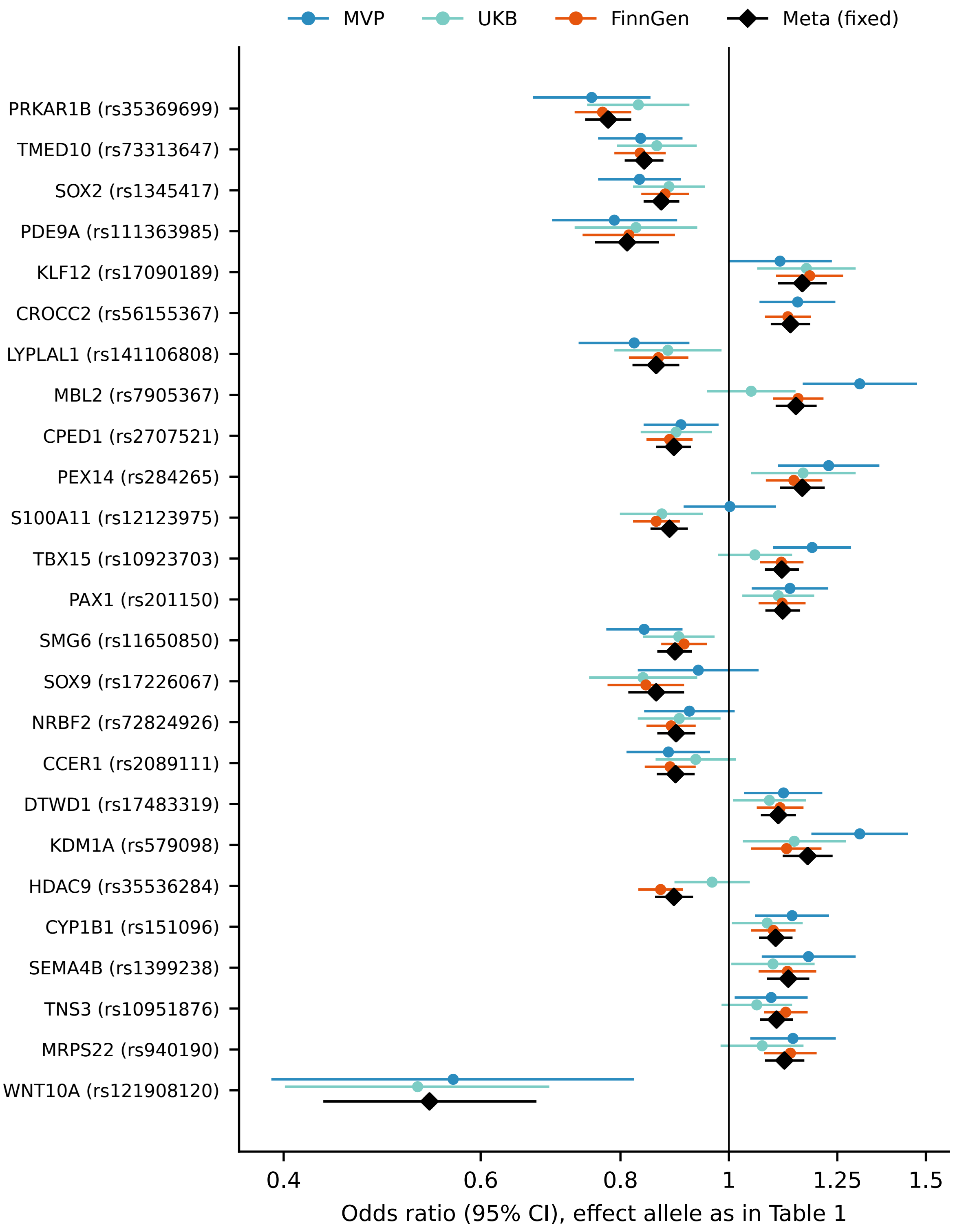


**Figure S2. Per-cohort effects at the 25 lead variants** (META_EUR cohorts and the fixed-effect meta-analysis, odds ratios with 95% confidence intervals). Missing points indicate a lead variant not measured in that cohort. Between-cohort heterogeneity was significant at *DKK1/MBL2*, *S100A11/TCHH*, *KDM1A* and *HDAC9* (Table S2).

### Supplementary tables

Provided as `supplementary_tables.xlsx`:

| Table | Content |
|---|---|
| S1 | PSD cohorts and comparator traits: cases, controls, effective N, variant counts, $\lambda_{GC}$, case definition, genotyping or sequencing, ancestry assignment, association model, covariates, relatedness handling, cohort-level variant QC, sex and age as reported, sources |
| S2 | Lead variants: GRCh38 and GRCh37 positions, fixed-effect (METAL) and random-effects OR (95% CI), Cochran's Q, $I^2$ and per-cohort OR |
| S3 | Locus classification (known/novel), leave-one-cohort-out *p* values |
| S4 | Robustness: lead-variant *p* values after LDSC-intercept correction and effects outside FinnGen |
| S5 | Fine-mapping summary (ABF and SuSiE-RSS with UK Biobank LD, LD-mismatch diagnostics, SuSiE-RSS with 1000 Genomes LD as a sensitivity analysis) |
| S6 | Credible-set variants with VEP annotation (PIP ≥ 0.1, and coding variants with PIP ≥ 0.01) |
| S7 | LDSC heritability, intercepts and liability-scale estimates |
| S8 | LDSC genetic correlations |
| S9 | Genomic SEM parameter estimates |
| S10 | Colocalisation of PSD loci with HS, acne, balding, BMI and WHRadjBMI (PP.H4 ≥ 0.5) |
| S11 | LAVA local genetic correlations (non-MHC blocks) |
| S12 | eQTL colocalisation (PP.H4 ≥ 0.5) |
| S13 | Locus-to-gene evidence |
| S14 | MHC-free MAGMA gene-set analysis: top sets, robustness reruns and the same sets in the comparator traits |

### Supplementary note: random-effects sensitivity analysis

Under DerSimonian-Laird random effects, 17 (META_EUR) and 18 (META_ALL) of the 25 loci remained genome-wide significant. The loci affected are those with between-cohort heterogeneity, together with lead variants near the threshold (Table S2).
